# Contemporary Resource Utilization and Costs among Patients with Medically Managed Severe Aortic Stenosis: Results from a US National Claims Database

**DOI:** 10.1101/2025.02.26.25322986

**Authors:** Adam S. Vohra, Shannon M. E. Murphy, Christin Thompson, David J. Cohen

## Abstract

**Background:** Patients with severe aortic stenosis (AS) who are managed medically have a poor prognosis. No contemporary studies have examined the costs and resource utilization associated with medical management for severe AS.

**Methods:** We used data from the Optum Market Clarity Database, which links electronic health records (EHR) with claims cost and utilization data, to identify patients with documented severe AS between 2016 and 2020 who did not undergo aortic valve replacement (AVR) within one year following diagnosis. To adjust for comorbidities that may have influenced treatment decisions, medically managed patients were matched 1:1 with similar patients who underwent AVR. Outcomes included healthcare resource utilization and costs during the 4-year period following the initial diagnosis of severe AS. Unadjusted Fine and Gray competing risk models were used to estimate resource utilization, and the Bang/Tsiatis method was used to quantify utilization and cost outcomes while accounting for censoring.

**Results:** Over the study period, 6,892 patients presented with newly diagnosed severe AS, of which 3334 (48%) were managed medically and 2812 (41%) were able to be propensity matched with patients who underwent AVR. Over 4 years of follow-up, these patients experienced an average of 1.99 hospitalizations—1.33 of which were related to a cardiovascular condition. Total healthcare-related costs (including hospitalizations, outpatient care, and pharmacy services) were $126,638/patient, of which $56,032 were related to inpatient care, $31,603 were related to outpatient care, and $21,160 were for pharmacy services.

**Conclusions:** In contemporary practice, despite the availability of effective treatment, many patients with documented severe AS do not undergo AVR within the first year after diagnosis. These patients with severe AS who are managed medically experience high health-care related resource utilization and costs. Further research is needed to identify factors associated with lack of timely valve replacement and to address barriers to care for these patients.

**Clinical Perspective:** What is new?

- This study examines the utilization and cost associated with untreated severe aortic stenosis in a contemporary U.S. cohort.

What are the clinical implications?

- Medically-managed patients with severe aortic stenosis are frail, elderly, with multiple comorbid conditions and have high resource utilization and costs.
- Further research is needed to identify patients who may benefit from aortic valve replacement.

---

Aortic stenosis (AS) is one of the most common forms of valvular heart disease in North America and Europe.^1, 2^ Historically, patients with severe, symptomatic AS who were managed medically had a dismal prognosis with 3-year mortality rates ranging from 50-80%.^3, 4^ In the PARTNER 1B trial of patients at extreme surgical risk, the medically managed group with severe symptomatic AS had a 1-year mortality of 50%.^5^ In addition to these high mortality rates, studies from the pre-transcatheter aortic valve replacement (TAVR) era found that patients with severe AS who were managed medically experienced high rates of hospitalization and annual healthcare related costs of about $30,000/year.^6^

Many of these extreme and high risk patients are now managed with TAVR.^7^ Although TAVR is currently recommended across the full surgical risk spectrum for patients with severe, symptomatic AS in whom valve replacement is not considered futile^8^, a sizeable proportion of patients with a Class I indication for AVR do not undergo either TAVR or SAVR.^9, 10^ However, little is known about the long-term outcomes of these patients with medically managed severe AS since the introduction of TAVR. We therefore sought to evaluate the real-world outcomes and healthcare related costs for a contemporary cohort of patients with severe AS who were managed medically.

## METHODS

### Data Source

This retrospective study used Optum’s de-identified Market Clarity dataset, which links electronic health records (EHR) with claims utilization and cost data from 2007 through 2023. Market Clarity is a U.S.-based database that captures over 70 million patient lives.^11^ The EHR component contains clinical data, echocardiographic findings, laboratory results, vital signs and measurements, diagnoses, procedures and clinical notes. The claims component includes medical and pharmacy claims from UnitedHealthcare, as well as from several other insurers and clearinghouses and represents a broad range of commercial, Medicaid, and Medicare health plans. Claims span both inpatient and outpatient settings and contain data elements such as hospital admission and discharge dates, diagnoses, procedures, and costs. Inclusion of claims data provides the ability to track patients across healthcare systems and thus allows for the longitudinal evaluation of clinical and cost outcomes. Market Clarity also includes health plan enrollment information used to define periods of data availability. The Optum de-identified Market Clarity dataset is deidentified and HIPAA compliant; therefore, institutional review board approval was not required for this study.

### Study Population

The initial study population comprised patients ≥18 years old with newly diagnosed severe aortic stenosis (AS) between 2016 and 2020. Severe AS was identified based on echocardiographic data and was defined in accordance with the 2020 ACC/AHA Heart Valve Disease Guidelines as mean pressure gradient ≥40 mm Hg, aortic valve area <1.0 cm^2^, or peak velocity ≥4.0 m/s, as documented in the EHR, as well as a current or prior diagnosis code indicating AS.^8, 12^ Patients with documentation of severe AS or an aortic valve procedure prior to the study period were excluded. In addition to a confirmed diagnosis, patients had to have medical health plan coverage for 6 months before the index diagnosing encounter (to allow for retrospective identification of comorbidities). Finally, study patients must have survived the index encounter and had continued health plan coverage for at least one day during the post-diagnosis study period. Patients with metastatic cancer are generally not considered as candidates for aortic valve replacement given to their short life expectancy and thus were excluded from the study cohort.

Among all newly diagnosed severe AS patients, the study population was further restricted to patients who did not receive AVR within the first year following diagnosis but who were eligible for treatment. To assess treatment eligibility, untreated patients were propensity score matched to *treated* patients (i.e., those who received AVR in the first year) as a means of excluding untreated patients whose clinical characteristics suggest they were not potential candidates for AVR. Figure 1 summarizes the full attrition diagram for the study cohort, and the Supplemental Materials provide additional details regarding propensity score matching, and characteristics of the full (unmatched) cohort and the AVR cohort used for matching (Supplementary Table S1).

**Figure 1.**
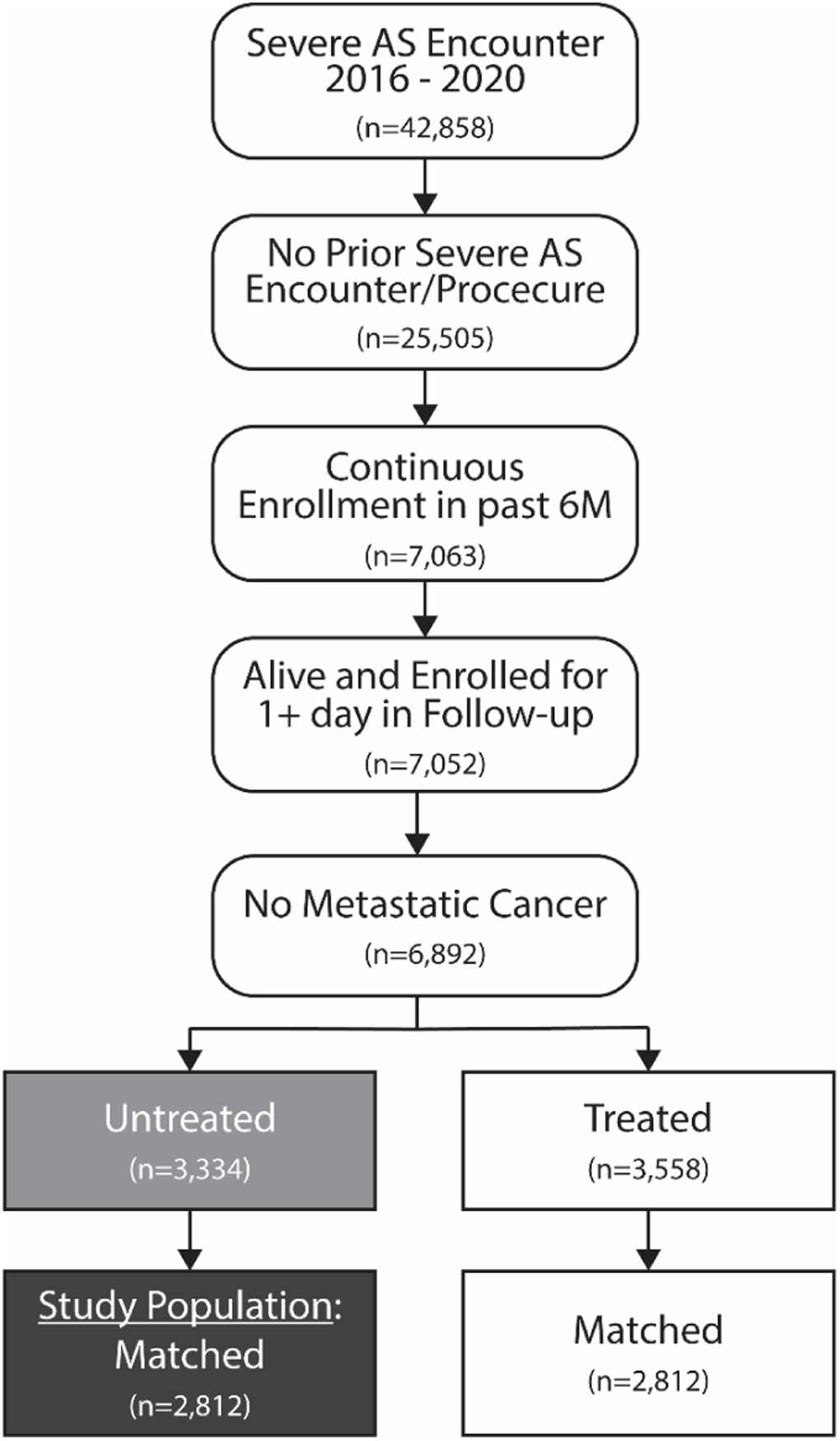
Attrition diagram for study population. After identification of patients with untreated severe AS, patients were propensity-matched with patients who underwent AVR in order to limit the study cohort to patients who were likely to be suitable candidates for AVR.

### Outcomes

Study outcomes included healthcare resource utilization and costs in the 4-year period following the initial diagnosis of severe AS. Cost in Market Clarity reflects standardized estimates of the allowed payment amounts (i.e., the insurance payment amounts plus the patient payment amounts). For this study, we examined costs both in aggregate and separately for inpatient, outpatient, and pharmacy services. Costs and utilization counts were also calculated for key drivers, including inpatient hospitalizations, skilled nursing facility days, cardiac rehabilitation days, and emergency department visits. Hospitalizations for cardiovascular conditions and heart failure were identified based on primary diagnosis codes. Outcomes were assessed for patients through 4 years following their first severe AS diagnosis, with censoring for death, health plan disenrollment, or receipt of AVR.

### Covariates

Patient demographic and clinical characteristics at the time of initial AS diagnosis were used to identify (via propensity score matching) and describe the study AS cohort. Characteristics included age, sex, race, payor, region, Elixhauser Comorbidity Score, Hospital Frailty Risk Score, diabetes, nonmetastatic cancer, dementia, heart failure, atrial fibrillation, chronic obstructive pulmonary disease, oxygen dependence, renal failure, end-stage renal disease on dialysis, prior stroke/transient ischemic attack, prior (non-aortic) valve procedure, bicuspid aortic valve disease, and presence of symptoms. Clinical characteristics were identified based on International Classification of Diseases, Ninth and Tenth Revisions (ICD-9 and 10), diagnosis and procedure codes (see Table S2 in the Supplemental Materials for a full list of diagnosis and procedure codes). The Elixhauser Comorbidity Score is a validated predictor of mortality based on 31 comorbidities, with scores ranging from 0 to 31 ^13^. The Hospital Frailty Risk Score uses ICD-10 diagnostic codes from claims documented in the prior year to estimate the risk of frailty and has been shown to predict mortality and other adverse health outcomes. Scores range from 0 to 45, with 0 to <5 considered low risk for frailty, 5 to 15 considered intermediate risk, and >15 to 45 considered high risk.^14, 15^

Symptomatic patients were identified based on either a heart failure diagnosis (per ICD- 10 code), ≥2 unique symptoms frequently associated with aortic stenosis, or the same symptom noted on multiple days, documented in the 6 months prior to or during the diagnosing encounter. Aortic stenosis symptoms were identified based on natural language processing of physician notes and included the following: chest pain or pressure, dyspnea with exertion, dyspnea at rest, and presyncope or syncope.^12^ Patients with missing sex (n=2, <0.1%), race (n=238, 3.5%) or region (n=186, 2.7%) were imputed with the mode (male, white and Midwest, respectively), and missing payor (n=338, 4.9%) was imputed as Commercial for patients age < 65 and Medicare for age ≥65.

### Statistical Analyses

Continuous variables are described as mean ± standard deviation, and categorical variables are described as counts and percentages. We used greedy 1:1 propensity score matching to patients with severe AS who underwent AVR in order to limit the untreated severe AS patients to those most likely to receive AVR. The propensity score was based on a logistic regression model to predict receipt of AVR within the year after the diagnosis of severe AS and included the aforementioned demographic and clinical covariates. All utilization and cost metrics were analyzed in two ways: 1) annually for patients who remained enrolled and untreated at the beginning of each year, and 2) cumulatively for the full study population. Resource utilization is reported both as any use of the health care service at the patient level and as the number of units of service utilized per patient (including patients with zero utilization). Patients were censored if they died, received AVR or left the health plan before the end of the 4-year follow-up period. The rate of any utilization in years 1 through 4 was estimated based on the cumulative incidence obtained via unadjusted Fine and Gray competing risks models for the patients alive and enrolled at the beginning of each year. For all other utilization and cost metrics, the Bang/Tsiatis method was used to account for censoring, and bootstrapping (1000 replicates) was used to calculate 95% confidence intervals.^16^

## RESULTS

### Patient Population

From the initial study population of 6,892 diagnosed severe AS patients from the Market Clarity Data, 3,334 (48%) were untreated in the year after diagnosis. After propensity matching with severe AS patients who underwent AVR in the year after diagnosis, the final study cohort consisted of 2,812 patients with severe, medically-managed severe AS.

Characteristics of the medically managed severe AS cohort suitable for AVR (n=2,812) are summarized in Table 1. The mean age was 74.1 years, 48% were female, 94% were Caucasian, and a majority were covered by Medicare. Based on EHR data, 70% of the cohort had symptoms consistent with AS. As compared with the full cohort of untreated patients, the portion considered suitable for AVR based on propensity score matching were slightly younger, more likely to be male and Caucasian, had lower Elixhauser and Hospital Frailty Risk Scores and lower rates of comorbidities such as dementia, atrial fibrillation, renal failure, and prior stroke. Supplemental Table S1 shows untreated and treated patient characteristics before and after matching.

**Table 1.** Baseline Patient Characteristics of the Untreated Severe AS Cohort.

| **Characteristic** | **Treatment-Eligible**<br>**(Matched) n=2,812** |
| --- | --- |
| Age in years, mean±SD | 74.1±11.6 |
| Female, % | 1,352 (48.1) |
| Race |  |
| Caucasian, % | 2,638 (93.8) |
| Black, % | 145 (5.2) |
| Asian, % | 29 (1.0) |
| Payor |  |
| Medicare, % | 1,604 (57.0) |
| Commercial, % | 1,072 (38.1) |
| Medicaid, % | 68 (2.4) |
| Other Payor, % | 34 (1.2) |
| Uninsured, % | 34 (1.2) |
| Region |  |
| Midwest, % | 1,495 (53.2) |
| Northeast, % | 537 (19.1) |
| South, % | 491 (17.5) |
| West, % | 289 (10.3) |
| Elixhauser score, mean±SD | 6.8±3.5 |
| Hospital Frailty Risk score, mean±SD | 9.7±9.5 |
| Comorbidities |  |
| Diabetes, % | 1,105 (39.3) |
| Cancer, % | 371 (13.2) |
| Dementia, % | 116 (4.1) |
| Heart failure, % | 1,160 (41.3) |
| Atrial fibrillation, %` | 899 (32.0) |
| COPD, % | 861 (30.6) |
| Oxygen dependence | 121 (4.3) |
| Renal failure, % | 827 (29.4) |
| On dialysis, % | 54 (1.9) |
| Prior stroke/TIA, % | 572 (20.3) |
| Prior non-aortic valve procedure, % | 4 (0.1) |
| Bicuspid aortic valve disease, % | 183 (6.5) |
| Symptomatic, % | 1,968 (70.0) |

TIA = transient ischemic attack; COPD = chronic obstructive pulmonary disease

### Clinical Outcomes and Resource Utilization

Of the 2,812 untreated patients in the study cohort, the following portion remained alive, enrolled, and untreated in the subsequent years: Year 2 (n=2,085, 74%), Year 3 (n=1,372, 49%), and Year 4 (n=926, 33%). The most common cause of study attrition during the 4-year follow-up period was death (n=837) followed by plan disenrollment (n=792) and receipt of AVR (n=620).

Among the study cohort of patients with untreated severe AS, the 4-year incidence of any hospitalization was 74%, and the incidence of any cardiovascular hospitalization and any heart failure hospitalization were 63% and 30%, respectively. When each hospitalization was counted separately, there was an average of 1.99 hospitalizations per patient over the 4-year follow-up period. Of these, 1.33 were cardiac-related and 0.51 were for heart failure. Additional resource use included 1.23 emergency room visits, 12.86 hospital days, and 9.00 skilled nursing facility days per patient (Table 2). The number of hospitalizations and hospital days were highest in the first year following the initial diagnosis of severe AS.

**Table 2.** Annual and Cumluative 4-Year Healthcare Resource Utilization.

| Utilization Measure | Year 1* | Year 2* | Year 3* | Year 4* | Cumulative (all patients)** |
| --- | --- | --- | --- | --- | --- |
| Sample Size* | 2,812 | 2,085 | 1,372 | 926 | 2,812 |
| Event incidence |  |  |  |  |  |
| Any hospitalizations, % | 45 (44 - 47) | 35 (32 - 37) | 32 (30 - 35) | 31 (28 - 35) | 74 (72 - 76) |
| CV hospitalizations, % | 36 (34 - 38) | 26 (25 - 28) | 25 (23 - 28) | 25 (23 - 29) | 63 (61 - 65) |
| HF hospitalizations, % | 15 (14 - 17) | 12 (10 - 13) | 10 (9 - 12) | 10 (8 - 12) | 30 (28 - 32) |
| SNF stay, % | 13 (12 - 14) | 10 (9 - 12) | 10 (9 - 11) | 12 (9 - 14) | 25 (23 - 27) |
| Cardiac rehab stay, % | 3 (3 - 4) | 1 (1 - 2) | 1 (1 - 2) | 0 (0 - 1) | 5 (4 - 6) |
| ED visit | 27 (26 - 29) | 27 (25 - 29) | 27 (24 - 29) | 29 (26 - 33) | 55 (53 - 57) |
| Total number of events |  |  |  |  |  |
| Any hospitalizations, | 0.91 (0.85 – 0.97) | 0.65 (0.58 – 0.71) | 0.57 (0.51 – 0.64) | 0.56 (0.48 – 0.64) | 1.99 (1.87 – 2.11) |
| CV hospitalization | 0.62 (0.57 – 0.66) | 0.42 (0.38 – 0.47) | 0.38 (0.33 – 0.43) | 0.37 (0.32 – 0.43) | 1.33 (1.24 – 1.41) |
| HF hospitalizations | 0.23 (0.21 – 0.26) | 0.17 (0.14 – 0.20) | 0.14 (0.12 – 0.17) | 0.14 (0.10 – 0.18) | 0.51 (0.46 – 0.56) |
| Resource |  |  |  |  |  |
| Hospital days | 5.82 (5.23 – 6.48) | 4.17 (3.62 – 4.74) | 3.56 (3.05 – 4.05) | 3.93 (3.22 – 4.81) | 12.86 (11.86 – 13.89) |
| SNF days | 3.02 (2.47 – 3.63) | 3.14 (2.46 – 3.88) | 3.31 (1.99 – 5.29) | 3.67 (2.53 – 5.03) | 9.00 (7.48 – 10.78) |
| Cardiac rehab days | 0.62 (0.48 – 0.78) | 0.19 (0.12 – 0.28) | 0.29 (0.14 – 0.44) | 0.07 (0 – 0.18) | 0.97 (0.78 – 1.17) |
| ED visits | 0.47 (0.43 – 0.51) | 0.42 (0.37 – 0.47) | 0.41 (0.35 – 0.47) | 0.45 (0.39 – 0.52) | 1.23 (1.12 – 1.33) |

Values in parentheses represent 95% confidence intervals; SNF = skilled nursing facility; ED = emergency department; HF = heart failure; CV = cardiovascular
\*Annual and cumulative event and resource counts are adjusted for censoring using the Bang-Tsiatis method<sup>16</sup>
\*\*Cumulative event and resource counts are less than the sum of annual counts due to retention of patients who died during follow-up

### Costs

Figure 2 displays the annual and cumulative costs for the study cohort over the 4 years following the initial diagnosis of severe AS. After accounting for censoring, total 4-year costs were $126,638 (including patients who died). Costs were highest in year 1 ($50,395) but remained >$40,000/surviving patient in years 2-4 as well. Table 3 summarizes annual and cumulative costs by category. The largest contributor to total 4-year costs were hospitalization costs ($56,032), while outpatient services and pharmacy services accounted for $31,603 and $21,160 per patient, respectively.

**Figure 2.**
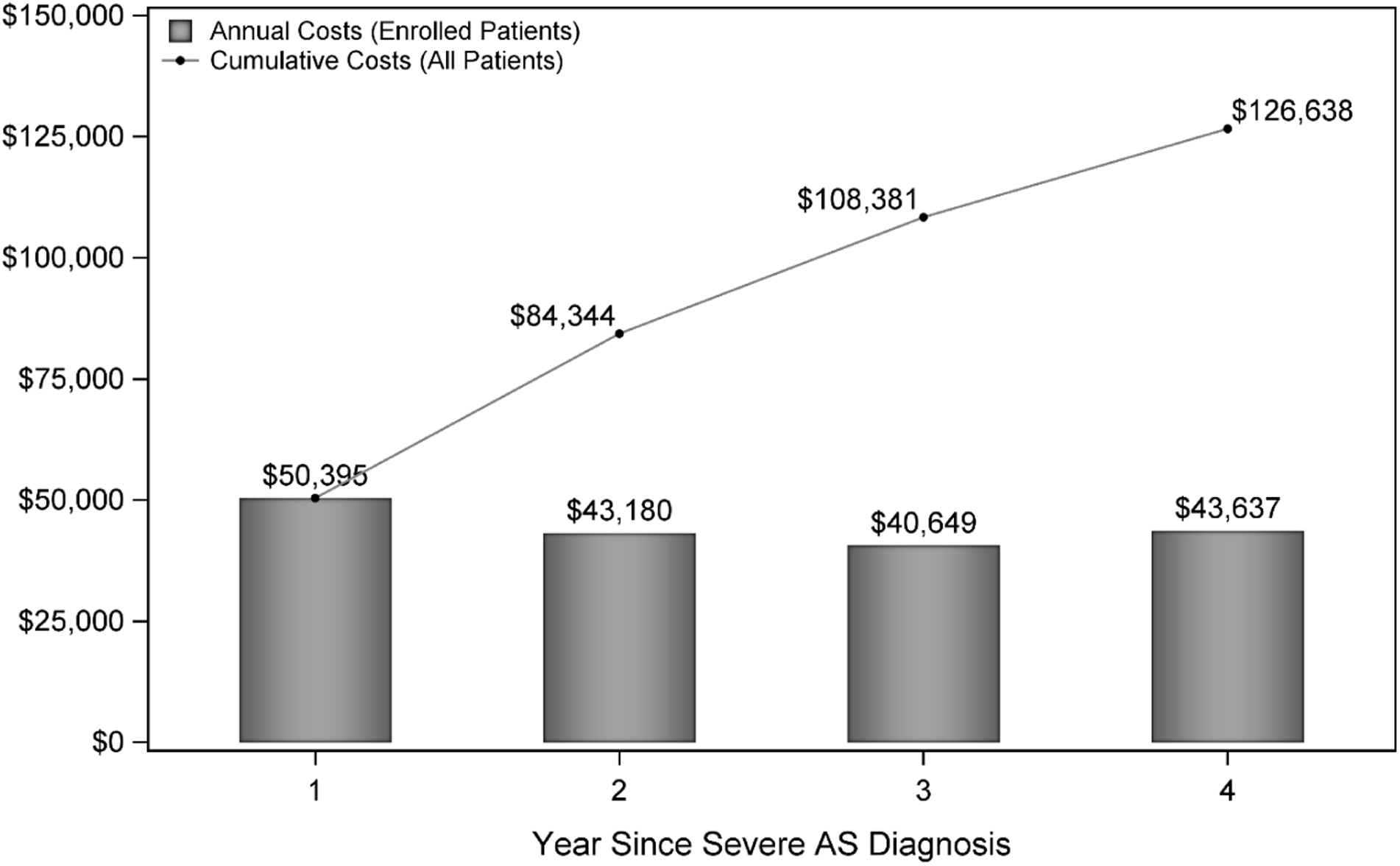
Annual and Cumulative Costs over 4 years after Diagnosis of Severe AS. Cumulative costs per patient are less than the sum of annual costs because of inclusion of zero costs after death for patients who died. AS = aortic stenosis

**Table 3.** Annual and Cumulative 4-year Costs.

| **Resource Category** | **Year 1**<br>**(n=2,812)** | **Year 2**<br>**(n=2,055)** | **Year 3**<br>**(n=1,372)** | **Year 4**<br>**(n=926)** | **Cumulative**<br>**(n=2,812)\*** |
| --- | --- | --- | --- | --- | --- |
| Total Cost<br>(Medical + Pharmacy) | \$50,395<br>(\$47,503 - \$53,377) | \$43,180<br>(\$39,675 - \$46,824) | \$40,649<br>(\$37,148 - \$44,572) | \$43,637<br>(\$38,788 - \$48,599) | \$126,638<br>(\$119,954 - \$133,341) |
| Inpatient Hospitalization<br>Costs | \$24,071<br>(\$22,008 - \$26,357) | \$18,837<br>(\$16,449 - \$21,408) | \$17,159<br>(\$14,539 - \$19,875) | \$16,754<br>(\$14,030 - \$19,699) | \$56,032<br>(\$51,884 - \$60,101) |
| CV Hospitalizations | \$20,279<br>(\$18,404 - \$22,306) | \$15,205<br>(\$13,024 - \$17,707) | \$13,671<br>(\$11,361 - \$16,160) | \$12,766<br>(\$10,408 - \$15,337) | \$45,656<br>(\$41,896 - \$49,580) |
| HF Hospitalizations | \$9,021<br>(\$7,622 - \$10,549) | \$7,426<br>(\$5,747 - \$9,484) | \$5,825<br>(\$4,130 - \$7,866) | \$4,813<br>(\$3,277 - \$6,473) | \$20,334<br>(\$17,776 - \$23,324) |
| SNF stays | \$2,196<br>(\$1,808 - \$2,627) | \$2,342<br>(\$1,615 - \$3,282) | \$1,788<br>(\$1,213 - \$2,434) | \$3,096<br>(\$1,636 - \$4,909) | \$6,389<br>(\$5,299 - \$7,655) |
| Cardiac Rehab | \$929<br>(\$584 - \$1,361) | \$467<br>(\$186 - \$814) | \$328<br>(\$144 - \$578) | \$81<br>(\$3 - \$210) | \$1,524<br>(\$1,060 - \$2,048) |
| ED visits | \$590<br>(\$536 - \$649) | \$547<br>(\$485 - \$613) | \$504<br>(\$438 - \$570) | \$664<br>(\$529 - \$857) | \$1,598<br>(\$1,468 - \$1,733) |
| Outpatient Medical Services | \$12,482<br>(\$11,646 - \$13,382) | \$10,442<br>(\$9,589 - \$11,357) | \$10,429<br>(\$9,384 - \$11,598) | \$11,327<br>(\$9,837 - \$12,789) | \$31,603<br>(\$29,694 - \$33,539) |
| Pharmacy Costs (outpatient) | \$7,176<br>(\$6,254 - \$8,317) | \$7,756<br>(\$6,579 - \$9,025) | \$7,542<br>(\$6,467 - \$8,806) | \$8,197<br>(\$6,551 - \$10,098) | \$21,160<br>(\$18,750 - \$23,950) |

Values in parentheses represent 95% confidence intervals; SNF = skilled nursing facility; ED = emergency department; CV = cardiovascular; HF = heart failure
\* Cumulative totals will not equal the sum of the totals in Years 1 - 4 due to the inclusion of patients who died
\*\* 95% confidence interval based on bootstrapping with 1000 samples

## DISCUSSION

In this study of commercially-insured patients with severe aortic stenosis in the TAVR era, we found that nearly half of these patients were managed conservatively—at least over the first year of follow-up. While these rates of medical management may appear high, they are comparable to those seen by others in a large, academic healthcare system.^17^ Although 22% of the medically managed cohort eventually underwent AVR over the 4-year follow-up period, the vast majority never underwent valve replacement. In addition, we found that resource utilization and costs for this population were high with mean 4-year costs of > $125,000 per patient and >$40,000 per year among surviving patients. These costs were driven predominantly by cardiovascular hospitalizations (mean 4-year cost $46,656/patient) as well as by outpatient health care services (mean 4-year cost $31,603/patient). Our findings of high annual costs among surviving patients are similar to those from Medicare beneficiaries during the pre-TAVR era ($29,278/year) ^6^ as well as those from the control group of inoperable patients studied in the PARTNER 1B trial ($53,621/year. ^18^ Our study extends these historical data by demonstrating that the high cost of patients with medically-managed severe AS continues to apply in the TAVR era.

In addition to demonstrating the high cost of medical management for patients with severe AS, our study also provides insight into the types of patients with severe AS who do not undergo AVR in the TAVR era. Not surprisingly, patients with severe AS who do not undergo prompt valve replacement tend to be elderly, frail, and have a high number of comorbid conditions including COPD, chronic kidney disease, and prior stroke or TIA. These findings suggest that many of the patients who are medically treated may have prognosis-limiting comorbid conditions that may have influenced treatment decisions. While it is intuitive that such patients would derive less survival benefit from AVR than patients without comorbidities, these patients are similar (albeit about 10 years younger) to patients enrolled in the original PARTNER 1B trial.^5^ In that trial, TAVR was found to reduce 1-year mortality by ∼50% and to increase quality-adjusted life expectancy by ∼1.3 years. Although projected lifetime costs remained substantially higher with TAVR, the incremental cost-effectiveness ratio of $50,000/QALY gained represents high economic value within the US healthcare system^18^ and is likely to be even more favorable (i.e., lower) in current practice given reductions in both procedural and hospital resource utilization over the past decade.^19–21^

Taken together with previous research, our findings thus suggest that even in the TAVR era, there is a large population of patients with severe aortic stenosis who are currently being managed conservatively and who may derive substantial benefit from either transcatheter or surgical AVR. Continued efforts to educate both clinicians and patients about the availability and health benefits of these therapies and to overcome well-recognized care disparities^22^ are thus warranted.

### Limitations

Our study has several limitations. First, we used data from a combination of insurance claims and electronic health records to identify patients with evidence of severe aortic stenosis who were managed medically but who likely could have been treated with TAVR. While we used propensity-score matching to limit the study cohort to medically managed patients who were similar to those who underwent AVR, our matching process could not account for unmeasured factors that may provide a valid rationale for conservative management (e.g., extreme frailty, unfavorable valve or vascular anatomy). In addition, although 70% of the study cohort had symptoms consistent with severe aortic stenosis, in some cases, these symptoms may have been attributed to other conditions such as chronic lung disease or heart failure with reduced ejection fraction. Second, given limitations in the claims dataset, it is likely that some deaths during follow-up may have been missed. Finally, while the Market Clarity dataset provided significant advantages such as longitudinal outpatient and inpatient follow-up, access to data on commercially-insured patients, and the ability to extract text-based data from the EHR using natural language processing, this dataset of patients—all of whom had commercial health insurance (including Medicare Advantage) may not be completely representative of all patients with severe AS.

### Conclusions

In conclusion, in a contemporary population of insured patients in an era when TAVR is widely available in the US, nearly half of all patients with newly diagnosed severe aortic stenosis are treated conservatively—at least for the first year of follow-up. These patients tend to be elderly, have multiple comorbid conditions, and experience relatively high rates of mortality, rehospitalization, and incur high annual and cumulative healthcare costs. Given the known benefits of both surgical and transcatheter aortic valve replacement for this population, efforts to improve detection and recognition of severe, symptomatic AS are warranted to ensure that all patients who may benefit from AVR are offered these therapies.

## Data Availability

he data that support the findings of this study are available from the corresponding author upon reasonable request.

## Acknowledgements

None.

## Nonstandard Abbreviations and Acronyms

AVR: aortic valve replacement
AS: aortic stenosis
ACC: American College of Cardiology
AHA: American Heart Association

